# Intermediate rectal dose is associated with late toxicity after prostate SBRT: a dose-volume histogram principal component analysis

**DOI:** 10.64898/2025.12.27.25343035

**Authors:** Amadeo J. Wals Zurita, Ana Illescas Vacas, Héctor Miras del Río, María Rubio Jiménez, Paula Vicente Ruíz, Jonathan Saavedra Bejarano, Francisco Carrasco Peña, Antonio Ureña, Mónica Ortiz Seidel

## Abstract

**Purpose:** To evaluate whether principal component analysis (PCA) of rectal and bladder dose-volume histograms (DVHs) identifies dose regions associated with late toxicity after prostate stereotactic body radiotherapy (SBRT).

**Methods/Patients:** This retrospective single-institution study included 106 patients with complete linked clinical and dosimetric data after prostate SBRT to 36.25 Gy in five fractions delivered every other day. PCA was applied separately to whole-organ and wall-based rectal and bladder DVHs. Associations with late grade >=2 gastrointestinal (GI) and genitourinary (GU) toxicity at 12 months were explored using Spearman correlation, ROC analysis and deliberately limited logistic regression.

**Results:** Among the 84 patients with available 12-month follow-up in the DVH-linked dosimetric cohort, grade >=2 toxicity was uncommon, with 3 GI events (3.6%) and 2 GU events (2.4%). Intermediate-dose rectal metrics showed the strongest exploratory signal for late GI toxicity, particularly rectal V18.1 Gy (AUC 0.868, 95% CI 0.740-0.997) and V29 Gy (AUC 0.827). These findings should be interpreted cautiously given the low number of events. Rectal wall metrics were less consistent, and no bladder or bladder-wall metric showed clinically meaningful discrimination for GU toxicity.

**Conclusions:** In this low-event SBRT cohort, intermediate whole-rectum dose was associated with late GI toxicity, but the findings are exploratory and require external validation. No isolated dosimetric predictor of late GU toxicity was identified.

## INTRODUCTION

Prostate cancer is a major indication for curative radiotherapy. Advances in intensity-modulated radiotherapy, image-guided radiotherapy and stereotactic body radiotherapy (SBRT) have enabled highly conformal dose distributions with improved sparing of surrounding organs at risk, particularly the rectum and bladder.

Hypofractionation in prostate cancer is supported by the low alpha/beta ratio of prostate adenocarcinoma. Multiple randomised phase III trials have demonstrated the non-inferiority of hypofractionated schedules compared with conventional fractionation, and five-fraction SBRT is now supported by contemporary randomised data, including PACE-B, with low rates of clinically significant late genitourinary (GU) and gastrointestinal (GI) toxicity [1–5]. Large multi-institutional series and meta-analyses have also reported favourable long-term outcomes [6,7].

Traditional dose-volume, normal-tissue complication probability and spatial dose-distribution approaches have shown that radiation toxicity may depend on both the magnitude and distribution of dose across organs at risk, while PCA-based analyses offer a way to address collinearity among DVH metrics and summarise global DVH shape [8–17].

Recent SBRT analyses suggest that toxicity prediction differs by organ. GI toxicity appears to follow dose-volume relationships, whereas GU toxicity may be more strongly associated with baseline urinary function and early treatment-related symptoms than with dosimetric parameters alone [18,19]. These findings suggest that conventional dosimetric approaches may not fully characterise toxicity risk across different organs at risk.

## MATERIALS AND METHODS

### Study design and patient population

This retrospective observational study analysed dose-volume histogram (DVH) patterns in organs at risk following stereotactic body radiotherapy (SBRT) for localised prostate cancer. Consecutive patients treated at a tertiary referral Radiation Oncology centre were screened.

Eligible patients had histologically confirmed localised prostate adenocarcinoma treated according to the institutional prostate SBRT protocol, which allowed cT1-cT3a N0 M0 disease. Patients were classified according to NCCN risk group. The primary analytical cohort was restricted to patients treated with a homogeneous SBRT regimen of 36.25 Gy delivered in five fractions of 7.25 Gy every other day, in order to preserve DVH comparability across patients. Patients with incomplete DVH data for rectum or bladder were excluded.

Institutional exclusion criteria for prostate SBRT included previous transurethral resection of the prostate or other prostate surgery, baseline IPSS >=19, disease beyond cT3a, nodal or metastatic disease, previous pelvic radiotherapy, hip prosthesis or osteosynthesis precluding adequate planning or image guidance, and clinically significant active anorectal pathology, including acute haemorrhoidal syndrome or active anal fissure.

The study flow is illustrated in Figure 1. From an initial cohort of 206 patients treated with prostate SBRT during the study period, DVH data were available for 176 patients. Among these, only patients treated with the homogeneous 36.25 Gy in 5 fractions regimen were included in the primary analysis. Although contemporary protocols such as PACE-B incorporated a simultaneous integrated boost up to 40 Gy to the CTV, the present cohort was restricted to patients treated with a homogeneous 36.25 Gy in five fractions regimen following an institutional urethra-sparing strategy based on the Novalis Circle Phase II approach. This restriction was applied to preserve DVH comparability across patients and prioritise reduction of genitourinary toxicity. After exclusion of cases with incomplete linkage between dosimetric and clinical data, the final analytical cohort consisted of 106 patients. These exclusions are important for interpretation, because they reduce sample size and limit generalisability; a sensitivity analysis including inhomogeneous boost schedules remains pending if the underlying dose data can be harmonised.

**Figure 1:**
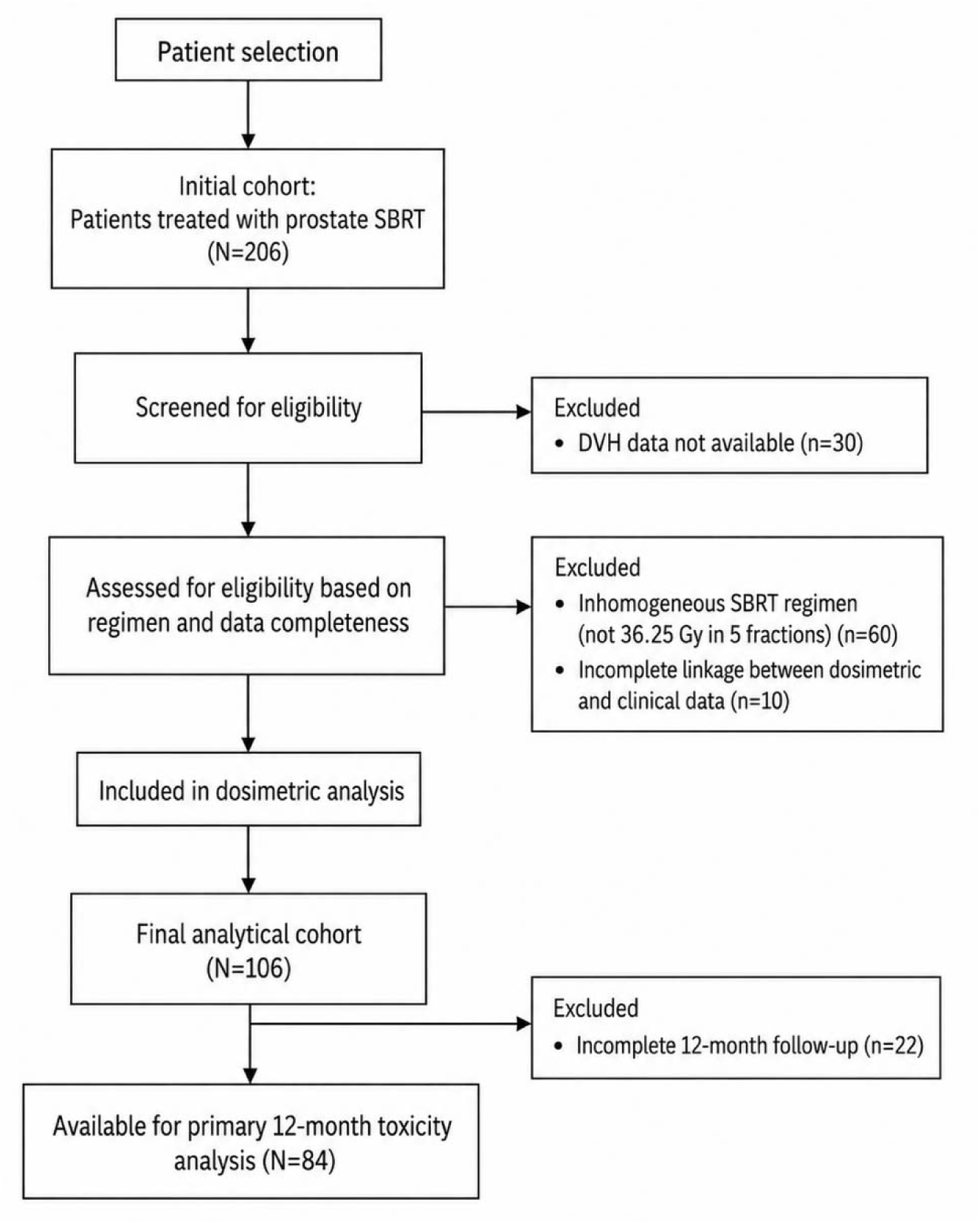
Flow diagram of patient selection.

Patient attrition reflected three independent sources of incomplete data inherent to the retrospective design: absence of DVH records in the institutional database (n=30), treatment with inhomogeneous SBRT regimens incompatible with DVH comparability (n=60), and incomplete linkage between dosimetric and clinical datasets (n=10). The subsequent reduction to 84 patients for the 12-month toxicity analysis reflects follow-up availability at that time point and does not represent an additional dosimetric exclusion. No formal comparison between included and excluded patients was performed, which should be considered a limitation.

Baseline clinical characteristics of the overall cohort are provided for context, while analyses were conducted on the subset with complete data.

Baseline clinical, tumour and volumetric characteristics of the cohort, including age, ECOG performance status, NCCN risk group, ISUP grade, baseline International Prostate Symptom Score (IPSS), androgen deprivation therapy use, and organ volumes, are summarised in **Table 1**. ADT duration followed institutional risk-adapted protocols, generally consisting of short-term ADT (4-6 months) for unfavourable intermediate-risk disease and long-term ADT (18-36 months, usually 24 months) for high-risk disease.

**Table 1.**
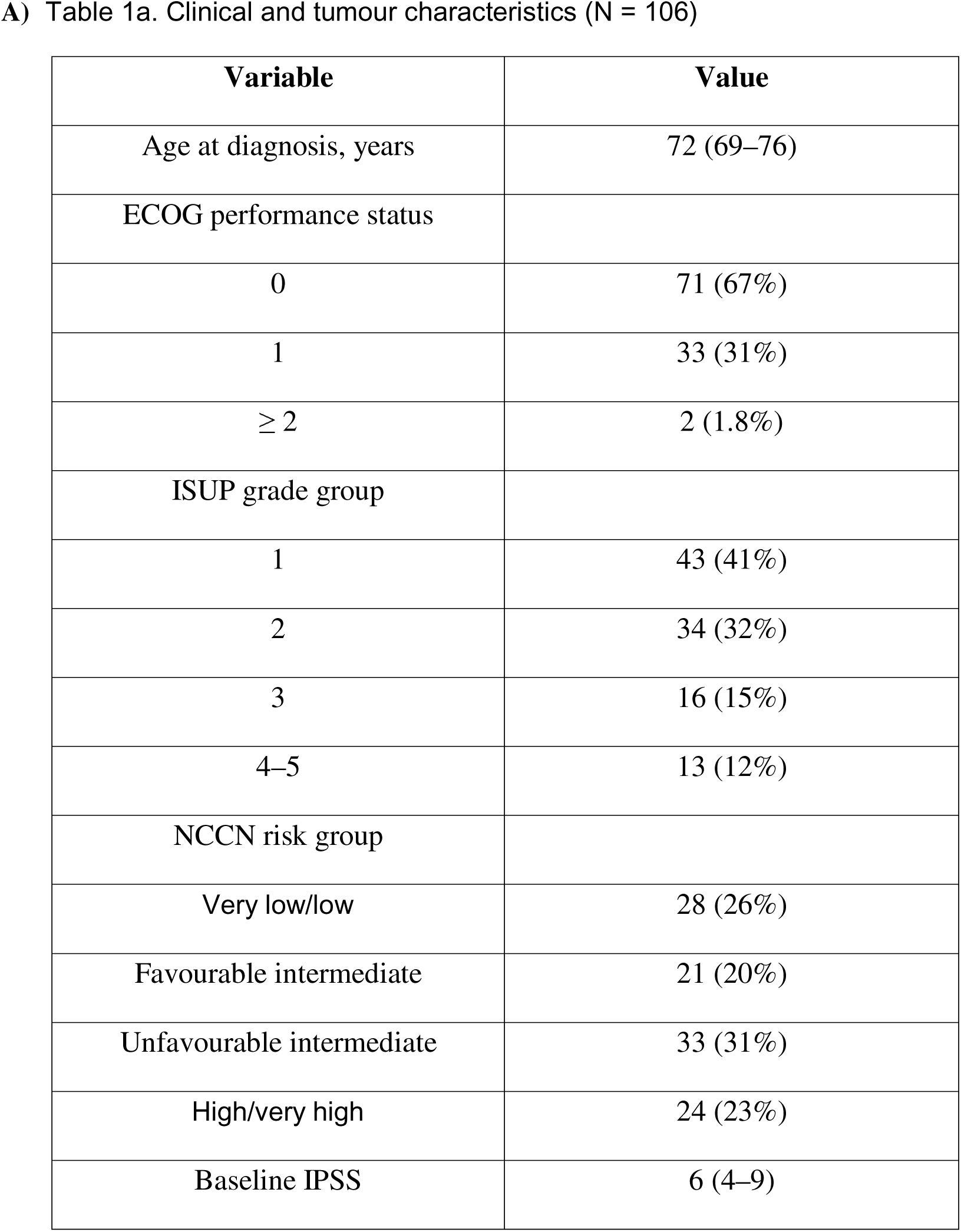
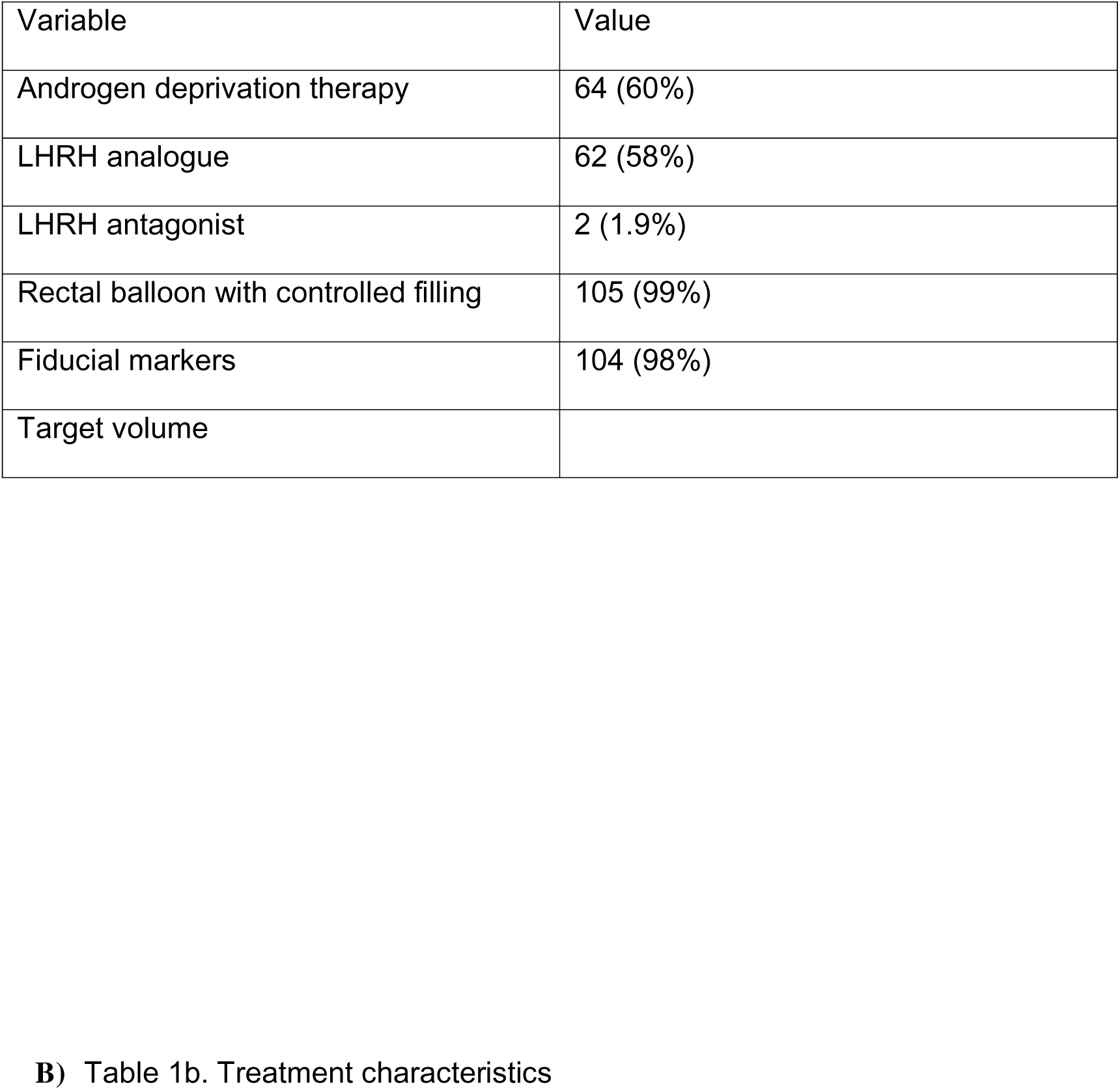
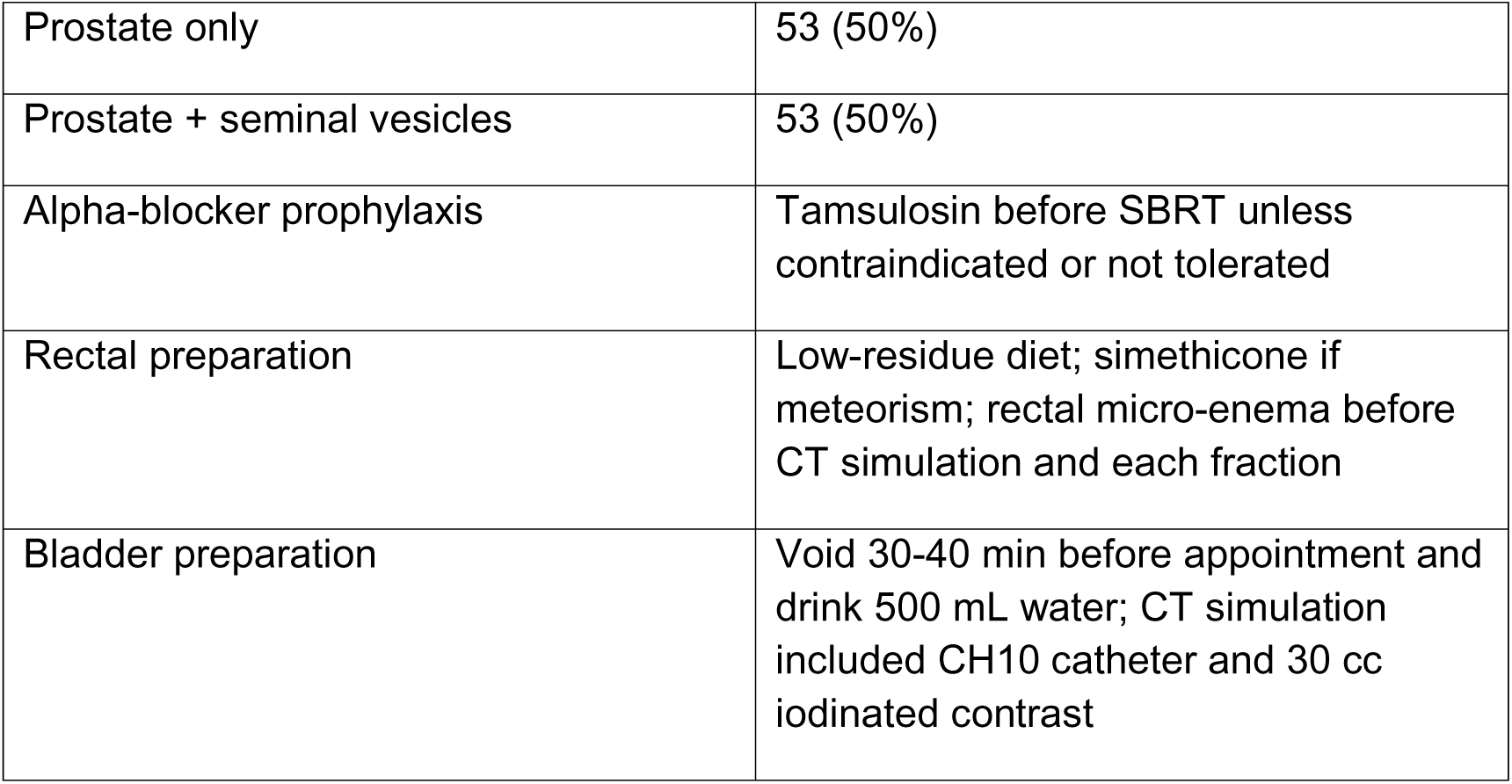
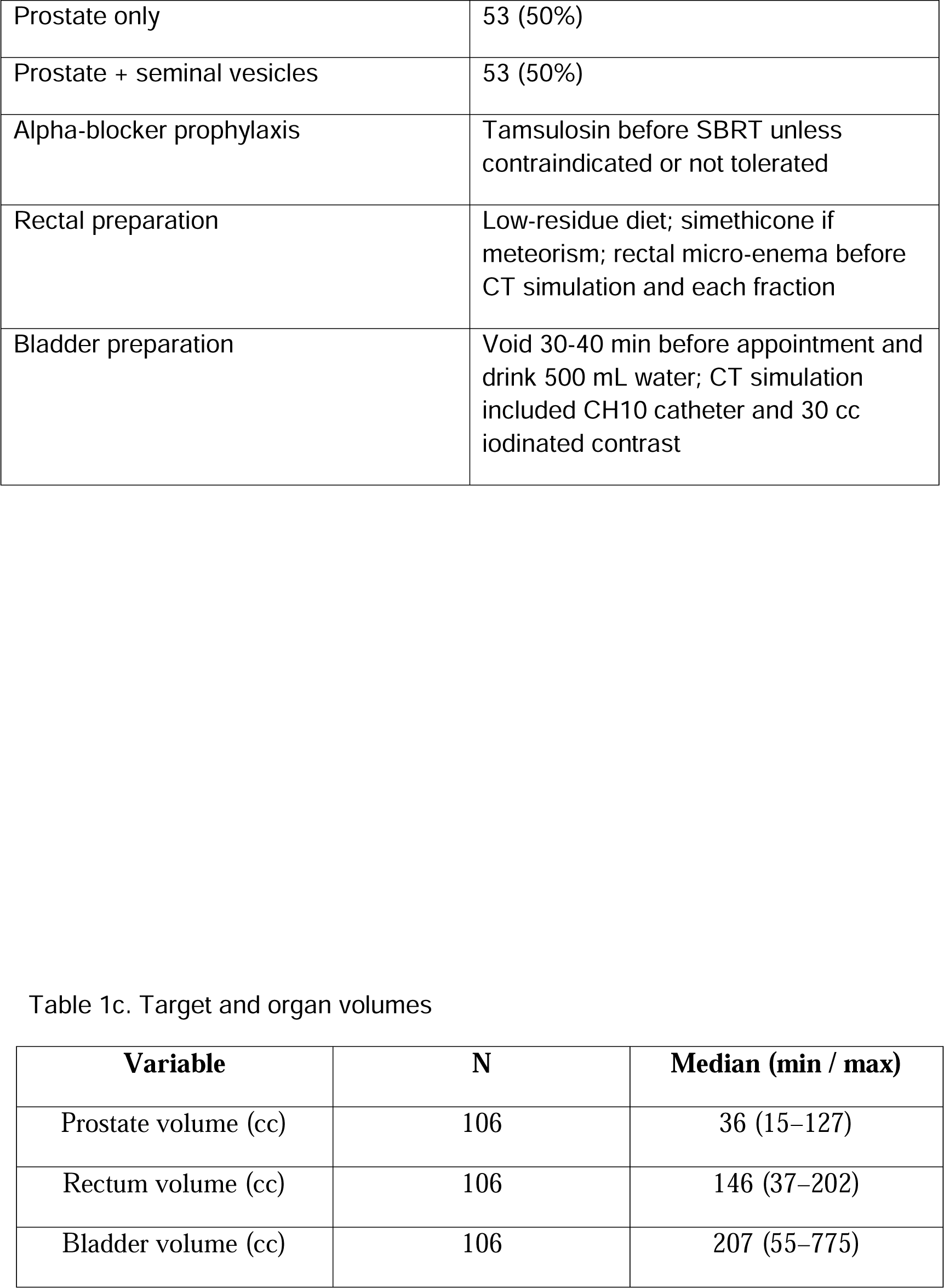
Baseline clinical, tumour and volumetric characteristics. Baseline characteristics of the study cohort. Continuous variables are reported as median (interquartile range), categorical variables as n (%).

### Treatment planning and delivery

Treatment planning followed the institutional prostate SBRT protocol, based on the prospective Novalis Circle Phase II approach. Plans were prescribed as 36.25 Gy in five fractions to the PTV, delivered on alternate days over a maximum of two weeks, with a urethra-sparing planning objective of 32.5 Gy in five fractions to the urethral planning volume. PTV coverage objectives included D98% >=95% of the prescription dose and D2% <=107%, with D2% <=110% considered acceptable. Organ-at-risk constraints included rectal wall V100% <5%, V90% <10% optimal (<15% acceptable) and V80% <20% optimal (<25% acceptable); bladder wall V100% <10-15%, V90% <20% and V50% <50%; femoral head D2% <=50%; and penile bulb mean dose <75% of the prescription dose.

The clinical target volume (CTV) included the prostate alone when the estimated risk of seminal vesicle involvement was <15% according to the Roach formula, and the prostate plus proximal seminal vesicles when this risk was >=15%. A planning target volume (PTV) was generated using a 5 mm isotropic expansion, reduced to 3 mm posteriorly.

The prescription dose was 36.25 Gy delivered to the PTV. This objective criterion is consistent with the balanced distribution of target volumes reported in Table 1, where the CTV included prostate alone in 50% of patients and prostate plus seminal vesicles in 50%.

Image guidance included intraprostatic fiducial markers and cone-beam CT. When fiducials were used, two VISICOIL™ fiducial markers were inserted transrectally under ultrasound guidance at least 7 days before CT simulation, whenever feasible.

Rectal immobilisation was achieved using a rectal balloon with controlled filling in the vast majority of patients, and no hydrogel rectal spacer was used. The institutional preparation protocol included a low-residue diet, simethicone when meteorism was reported, and a rectal micro-enema on the evening before and again before CT simulation or each treatment fraction. The endorectal balloon was filled with a standard 80 cc of air, with insertion depth adapted to reproduce the simulation position.

Bladder preparation consisted of voiding 30-40 minutes before the appointment followed by intake of 500 mL of water. At CT simulation, urethral catheterisation with a CH10 instillation catheter was performed and 30 cc of iodinated contrast was instilled; urethral catheterisation was not routinely required during treatment fractions. Prophylactic alpha-blocker therapy, mainly tamsulosin, was commonly initiated before SBRT unless contraindicated or not tolerated.

DVH data were standardised using Z-score normalisation. The number of components retained was determined primarily using parallel analysis, with the empirical Kaiser criterion (eigenvalue >1) used as a supportive rule for interpretability. Varimax rotation was applied to improve interpretability.

PCA outputs included eigenvalues, explained variance and dose regions associated with maximum component loadings. For each retained component, the dose corresponding to the maximum loading was identified, allowing localisation of the dose levels contributing most to inter-patient variability and facilitating clinical interpretation. Results are reported in Supplementary Table S2.

### Statistical analysis

Late toxicity was scored according to CTCAE v5.0. Associations between dosimetric variables and toxicity were evaluated using Spearman correlation, receiver operating characteristic (ROC) analysis, and binary logistic regression.

Dosimetric parameters were analysed as continuous variables. ROC analyses were used to evaluate discriminative performance using area under the curve (AUC).

Given the limited number of toxicity events, regression analyses were treated as exploratory signal detection. Multivariable estimates were attempted only as sparse-event sensitivity analyses and should not be interpreted as stable effect-size estimates. Extended analyses are detailed in Supplementary Table S5.

### Ethics

The study was approved by an institutional research ethics committee. All data were retrospectively collected, pseudonymised prior to analysis, and handled in accordance with applicable data protection regulations and the Declaration of Helsinki.

## RESULTS

A total of 106 patients with localised prostate cancer treated with stereotactic body radiotherapy with complete DVH data were included in the final dosimetric analysis. Baseline clinical and tumour characteristics are summarised in Table 1. The median age at diagnosis was 72 years (interquartile range [IQR]: 69–76). Most patients had a good performance status (ECOG 0–1 in 98%), and the cohort included a balanced distribution of NCCN risk groups, with low-risk disease in 26%, favourable intermediate-risk in 20%, unfavourable intermediate-risk in 31%, and high/very high-risk in 23%. The median baseline International Prostate Symptom Score (IPSS) was 6 (IQR: 4–9). Androgen deprivation therapy was administered in 60% of patients.

Regarding treatment characteristics, all patients received a homogeneous SBRT regimen of 36.25 Gy in five fractions. The clinical target volume included the prostate alone in 50% of cases and prostate plus seminal vesicles in the remaining 50%. The median prostate volume was 36.3 cc (IQR: 27.5–47.9). Rectal immobilisation was achieved using a rectal balloon in 99% of patients, and fiducial markers were used for image guidance in 98%. The rectal volume range in Table 1 should be interpreted in this context: although the institutional protocol used a standard 80 cc endorectal balloon, one patient in the analytical cohort was treated without a rectal balloon, consistent with the reported use of rectal immobilisation in 105 of 106 patients.

### Clinical outcomes

At 12 months after treatment, clinically relevant late toxicity (CTCAE v5.0 grade >=2) was infrequent. Among the 84 patients with available 12-month follow-up in the DVH-linked dosimetric cohort, the incidence of GU toxicity >= grade 2 was 2.4% (2 events), while GI toxicity >= grade 2 was 3.6% (3 events). The corresponding event phenotypes are shown in Table 2. These event counts and denominators should be considered when interpreting all exploratory analyses.

**Table 2.** Late toxicity at 12 months and event phenotype (N=84 with available 12-month follow-up; CTCAE v5.0)

| **Toxicity** | **Grade 0–1 n (%)** | **≥ Grade 2 n (%)** | **≥ Grade 2 phenotype** |
| --- | --- | --- | --- |
| Genitourinary (GU) | 82 (97.6%) | 2 (2.4%) | Urinary urgency (n=1); urinary tract obstruction (n=1) |
| Gastrointestinal (GI) | 81 (96.4%) | 3 (3.6%) | Constipation (n=1); rectal bleeding (n=2) |

Note: Phenotypes correspond to grade $\geq 2$ events at 12 months in the DVH-linked dosimetric cohort. DVH-linked denotes availability of valid dose-volume histogram data. One additional clinical-only GU event (urinary frequency; ID-200) lacked valid DVH data and was excluded from DVH-linked dosimetric analyses; see Supplementary Table S9.

The 12-month time point was selected for the primary toxicity analysis because it provided the most informative balance between event capture and cohort availability. As shown in Supplementary Table S1, grade >=2 GI events were observed up to 12 months but not beyond this time point in the grouped DVH-linked longitudinal table, whereas later follow-up included progressively fewer patients at risk. Accordingly, a 24-month analysis would have been less informative for rectal toxicity and more vulnerable to sparse-event instability.

Longitudinal analysis of the DVH-linked cohort showed that GU toxicity peaked at 24 months (6.9%) and subsequently declined, whereas GI toxicity remained low over time, with no DVH-linked GI grade >=2 events beyond 12 months in the grouped longitudinal table. One source-clinical grade 4 rectal bleeding event at 18 months occurred in a patient without valid DVH data and was therefore not included in the DVH-linked dosimetric analysis; this case is documented separately in Supplementary Table S8b. These results provide the clinical framework for the subsequent dosimetric analyses (Supplementary Table S1).

### Principal component analysis of DVHs

Principal component analysis demonstrated that a limited number of components explained the majority of inter-patient DVH variability across all analysed structures. For the whole rectum, the first component had its maximum loading in a high-dose region (42.5 Gy), whereas the second and third components localised to lower and intermediate dose levels (13.0 and 29.0 Gy). For the rectal wall, the first two components localised to 13.0 and 28.5 Gy. Thus, intermediate-dose regions contributed importantly to retained variability patterns, but they were not the sole dominant component across all rectal analyses.

This distinction is important for interpretation: the PCA loadings describe major sources of DVH variability, whereas the toxicity analyses identified which candidate metrics showed the strongest exploratory association with grade >=2 GI toxicity. The strongest toxicity signal was observed for whole-rectum V18.1 Gy, with a complementary signal at V29 Gy.

### Correlation analyses

Spearman correlation analyses demonstrated moderate correlations between selected low-to-intermediate rectal DVH metrics and planning target volume (PTV). The strongest correlation was observed for low-dose rectal V1.5 Gy (rho = 0.52), followed by V13 Gy (rho = 0.37) and V29 Gy (rho = 0.27; all p < 0.001). In contrast, high-dose rectal V42.5 Gy showed a weak, non-significant correlation with PTV (rho = 0.113), indicating that PTV coupling was not monotonic across the dose range.

A similar pattern was observed for bladder and bladder wall DVHs, where low-to-intermediate dose metrics were moderately correlated with PTV, whereas high-dose exposure demonstrated minimal or no correlation.

#### Predictive performance of DVH metrics

In receiver operating characteristic (ROC) analyses, rectal DVH metrics corresponding to intermediate-dose regions showed the highest exploratory discriminative performance for late rectal toxicity >= grade 2 at 12 months.

Among these, rectal V18.1 Gy demonstrated the strongest exploratory discriminatory ability (AUC = 0.868; 95% CI: 0.740-0.997), followed by V29 Gy (AUC = 0.827). This signal should be interpreted cautiously given the low number of events. Nevertheless, the emergence of V18.1 Gy as the most informative metric is clinically relevant because it supports the importance of an established intermediate-dose rectal constraint rather than proposing a new unvalidated cut-off. In contrast, low-dose (V1.5 Gy) and high-dose (V42.5 Gy) metrics showed poor or no discriminative performance.

Rectal wall metrics showed less consistent results. Although V13 Gy demonstrated moderate discrimination (AUC ≈ 0.78), its performance was less robust and associated with wider confidence intervals. No clinically meaningful predictive performance was observed for bladder or bladder wall DVH metrics, with AUC values consistently below 0.70.

#### Regression analyses

In univariable logistic regression models, rectal V18.1 Gy was significantly associated with late rectal toxicity >= grade 2 (p = 0.047). Because this endpoint included only 3 GI events at 12 months, all regression estimates are statistically fragile and should be interpreted as exploratory signal detection rather than reliable effect-size estimation. Additional model building was deliberately limited because strong collinearity among DVH metrics and sparse events precluded stable multivariable inference.

No significant associations were identified between bladder-related DVH metrics and genitourinary toxicity, and regression models for these endpoints showed limited stability.

The main exploratory dosimetric signals for late toxicity at 12 months are summarised in Table 3, highlighting an association between intermediate whole-rectum exposure and late GI toxicity, and the absence of clinically meaningful bladder or bladder-wall DVH signals for GU toxicity.

**Table 3.** Summary of exploratory dosimetric signals for late toxicity (12 months)

| Structure | DVH metric | Analysis | Key result | Interpretation |
| --- | --- | --- | --- | --- |
| Rectum | V18.1 Gy | ROC | AUC 0.868 | Exploratory discriminative signal |
| Rectum | V29 Gy | ROC | AUC 0.827 | Complementary exploratory signal |
| Rectum | V18.1 Gy | Logistic regression | p = 0.047 | Exploratory association; limited events |
| Rectal wall | V13 Gy | ROC | AUC 0.78 | Moderate, not robust |
| Bladder | Multiple | ROC | AUC < 0.70 | No discrimination |
| Bladder wall | Multiple | ROC | AUC < 0.70 | No discrimination |

Exploratory analyses of available non-dosimetric clinical variables, including age, prostate volume, baseline PSA and ISUP grade, did not identify stable independent associations with late rectal or genitourinary toxicity (Supplementary Table S6a and S6b). These analyses were limited by the small number of events, wide confidence intervals and incomplete availability of potentially relevant baseline functional variables.

Missing data reflect incomplete baseline documentation or unavailable imaging-derived volumes. No imputation was performed.

ADT duration followed institutional risk-adapted protocols, generally consisting of short-term ADT (4-6 months) for unfavourable intermediate-risk disease and long-term ADT (18-36 months, usually 24 months) for high-risk disease. Patient-level duration counts were not tabulated in this revision because ADT start/end dates were incomplete in a subset of patients and the applicable denominators varied according to DVH availability across rectal, rectal-wall, bladder and bladder-wall structures.

Rectal immobilisation was achieved using a rectal balloon with controlled filling; no hydrogel rectal spacer was used in this cohort. The rectal volume range should therefore be interpreted together with the recorded balloon-use variable: one patient in the analytical cohort was treated without a rectal balloon, which explains why the minimum contoured rectal volume is not constrained by the standard 80 cc balloon filling.

## DISCUSSION

This study provides an exploratory dosimetric analysis of late toxicity following prostate stereotactic body radiotherapy using a principal component analysis-based approach applied to dose-volume histograms. The main finding is that intermediate-dose exposure of the whole rectum was associated with clinically relevant late rectal toxicity, whereas no robust dosimetric predictors were identified for genitourinary toxicity.

These results are also supported by recent evidence from contemporary SBRT series, which have identified moderate-dose regions as key determinants of both gastrointestinal and genitourinary toxicity. In particular, Ozyigit et al. reported that intermediate-dose exposure was consistently associated with acute and late toxicity endpoints, reinforcing the relevance of this dose range across different clinical settings [19].

The near-universal use of rectal balloon immobilisation in this cohort may have contributed to the observed DVH patterns, particularly by reducing high-dose variability and emphasising intermediate-dose regions.

The predominance of intermediate-dose regions may reflect a volume-dependent component of rectal injury, whereby clinically relevant symptoms are influenced by the extent of irradiated rectal tissue. However, the rectum should not be considered a purely parallel organ, and focal high-dose exposure, spatial dose distribution, baseline bowel function and contouring variability may all contribute to toxicity risk. While high-dose constraints remain essential to prevent severe complications, our results suggest that intermediate-dose exposure across larger rectal volumes may still be relevant within accepted dose limits.

Traditional DVH metrics are highly correlated and may not fully reflect the overall dose distribution [8,9]. PCA allows a more global evaluation of dose patterns and may help identify clinically relevant dose regions associated with toxicity [13–15]. Spatial dose-distribution approaches similarly suggest that the location and extent of irradiated regions can add information beyond conventional DVH parameters [16,17].

In contrast to rectal toxicity, no clinically meaningful dosimetric predictors were identified for genitourinary toxicity. This negative finding should not be interpreted as evidence that planned dose distribution is irrelevant, because the low number of GU events limits statistical power. Rather, the present data did not demonstrate a stable bladder or bladder-wall DVH signal. Recent PACE-B analyses suggest that baseline urinary function and early treatment-related symptoms may be important predictors of late urinary toxicity, providing a plausible explanation for the absence of a strong DVH-only signal in this cohort [18].

This contrast between rectal and genitourinary toxicity highlights the organ-specific nature of radiation-induced toxicity. Rectal toxicity may include a clearer volume-dose component, whereas urinary toxicity after prostate SBRT probably reflects interactions among baseline urinary status, prostate size, urethral or bladder-neck dose, medication use, acute toxicity and clinical management. Future analyses should include baseline bowel function, haematuria or haematochezia, haemorrhoids, urinary medication use and other functional variables when available.

The homogeneous 36.25 Gy prescription used in this cohort should be interpreted in the context of the institutional urethra-sparing approach. While PACE-B incorporated a simultaneous integrated boost up to 40 Gy to the CTV, the present analysis deliberately excluded inhomogeneous boost schedules to maintain dosimetric comparability.

Therefore, the results should not be directly extrapolated to dose-escalated SBRT regimens. When interpreted in the context of landmark trials such as PACE-B, our findings should be considered complementary rather than contradictory. The every-other-day schedule used in the present cohort should also be interpreted in the context of PATRIOT, which compared every-other-day and once-weekly prostate SABR schedules [20]. Accordingly, extrapolation of the present dosimetric findings to once-weekly SBRT schedules should be made cautiously. The dose-volume constraints used in these trials were designed to ensure population-level safety and have proven effective in limiting severe toxicity. The restricted dose and planning range of this PACE-B-like cohort should therefore not be viewed solely as a limitation. Rather, it reflects contemporary SBRT practice, in which high-dose rectal constraints are routinely met and severe toxicity is uncommon. Within this clinically constrained setting, residual inter-patient variability is expected to occur predominantly in lower and intermediate-dose regions. The observation that rectal V18.1 Gy emerged as the most informative metric is therefore not unexpected, but it is clinically relevant: it supports the continued importance of intermediate-dose rectal sparing when evaluating otherwise acceptable plans. The present results support careful review of intermediate rectal dose during plan evaluation, not as a replacement for validated constraints and not as proof of a new threshold, but as independent, hypothesis-generating evidence that this already recognised dose region remains clinically meaningful in modern prostate SBRT.

Although target volume size may influence organ irradiation, its effect may be mediated partly through DVH parameters. Intermediate rectal dose metrics were moderately correlated with PTV volume, so the observed signal could reflect a mixture of target-size effects, anatomy and planning geometry. This limits causal interpretation and should be addressed in future validation cohorts.

Available non-dosimetric clinical variables did not demonstrate independent associations with toxicity in this cohort, but this should be interpreted cautiously. The analysis was underpowered for clinical predictors and did not include several baseline functional variables requested by reviewers, including baseline bowel symptoms, haemorrhoids/haematochezia and urinary medication use.

A conceptual summary of the differential dosimetric signal observed between rectal and bladder structures is provided in Supplementary Figure S3.

A summary of the key findings and their clinical implications is provided in Supplementary Table S7.

## CONCLUSIONS

In this institutional cohort of patients treated with prostate stereotactic body radiotherapy, clinically relevant late rectal toxicity was infrequent and comparable to rates reported in contemporary randomised trials. Despite the low number of events, exposure of the whole rectum to intermediate dose levels, particularly V18.1 Gy, showed an exploratory association with rectal toxicity >= grade 2.

Dosimetric parameters derived from the whole rectum outperformed those obtained from the rectal wall, suggesting that global volumetric dose burden may be more informative than focal wall metrics in this dataset. In contrast, no isolated dosimetric signals for late genitourinary toxicity were identified; this should be interpreted as absence of a demonstrable DVH signal rather than proof that urinary toxicity is independent of dose.

These findings complement population-based dose-volume constraints derived from landmark trials such as PACE-B and support the relevance of the established rectal V18.1 Gy intermediate-dose constraint during plan evaluation. PCA of DVHs may help identify clinically relevant dose regions, but these results remain supportive rather than definitive and require confirmation in larger cohorts.

## Data Availability

Anonymised clinical and dosimetric data underlying this study are available from the corresponding author upon reasonable request, subject to consultation with the institutional Research Ethics Committee and compliance with GDPR requirements.

## USE OF ARTIFICIAL INTELLIGENCE

A large language model (ChatGPT, OpenAI) was used as a writing and editorial support tool to assist with language refinement, structural editing, and consistency checking of the manuscript and supplementary materials. The model did not generate original data, perform statistical analyses, or influence study design, data interpretation, or scientific conclusions. All analyses, interpretations, and final content decisions were made by the authors.

## AUTHOR CONTRIBUTIONS

Conceptualisation: Amadeo J. Wals Zurita.

**Methodology:** Amadeo J. Wals Zurita, Ana Illescas Vacas.

**Formal analysis:** Amadeo J. Wals Zurita.

**Investigation:** Amadeo J. Wals Zurita, Ana Illescas Vacas, María Rubio Jiménez, Paula Vicente Ruíz, Héctor Miras del Río, Jonathan Saavedra Bejarano, Francisco Carrasco Peña, Antonio Ureña, Mónica Ortíz Seidel.

**Data curation:** Amadeo J. Wals Zurita, Héctor Miras del Río, Antonio Ureña, Mónica Ortíz Seidel.

**Writing – original draft:** Amadeo J. Wals Zurita.

**Writing – review & editing:** Ana Illescas Vacas, María Rubio Jiménez, Paula Vicente Ruíz, Héctor Miras del Río, Jonathan Saavedra Bejarano, Francisco Carrasco Peña, Antonio Ureña, Mónica Ortíz Seidel.

Visualisation: Amadeo J. Wals Zurita.

**Supervision:** Amadeo J. Wals Zurita, Ana Illescas Vacas.

**Project administration:** Amadeo J. Wals Zurita.

**Funding acquisition:** Not applicable.

*All authors have read and approved the final manuscript*.

## LIST OF ABBREVIATIONS

CBCT: Cone-beam computed tomography
CTCAE: Common Terminology Criteria for Adverse Events
CTV: Clinical target volume
DVH: Dose–volume histogram
GI: Gastrointestinal
GU: Genitourinary
NCCN: National Comprehensive Cancer Network
OAR: Organ at risk
PACE-B: Prostate Advances in Comparative Evidence – SBRT vs standard RT
PCA: Principal component analysis
PTV: Planning target volume
ROC: Receiver operating characteristic
SBRT: Stereotactic body radiotherapy

