## supplementary for "Intermediate rectal dose is associated with late toxicity after prostate SBRT: a dose-volume histogram principal component analysis": Supplementary_mayo_preprint_2026.docx

### Supplementary Appendix

#### Supplementary Table S1. Longitudinal evolution of genitourinary and gastrointestinal toxicity

Longitudinal incidence of genitourinary (GU) and gastrointestinal (GI) toxicity according to CTCAE v5.0 at predefined follow-up intervals (data derived from institutional analysis up to page 34).

| **Time point (months)** | **Patients at risk (n)** | **GU G0–1 n (%)** | **GU ≥G2 n (%)** |
| --- | --- | --- | --- |
| 6 | 96 | 92 (95.8%) | 4 (4.2%) |
| 12 | 84 | 82 (97.6%) | 2 (2.4%) |
| 18 | 65 | 63 (96.9%) | 2 (3.1%) |
| 24 | 58 | 54 (93.1%) | 4 (6.9%) |
| 30 | 40 | 40 (100.0%) | 0 (0.0%) |
| 36 | 27 | 27 (100.0%) | 0 (0.0%) |
| 42 | 6 | 6 (100.0%) | 0 (0.0%) |
| 48 | 1 | 1 (100.0%) | 0 (0.0%) |
| **Time point (months)** | **Patients at risk (n)** | **GI G0–1 n (%)** | **GI ≥G2 n (%)** |
| 6 | 96 | 94 (97.9%) | 2 (2.1%) |
| 12 | 84 | 81 (96.4%) | 3 (3.6%) |
| 18 | 65 | 65 (100.0%) | 0 (0.0%) |
| 24 | 58 | 58 (100.0%) | 0 (0.0%) |
| 30 | 40 | 40 (100.0%) | 0 (0.0%) |
| 36 | 27 | 27 (100.0%) | 0 (0.0%) |
| 42 | 6 | 6 (100.0%) | 0 (0.0%) |
| 48 | 1 | 1 (100.0%) | 0 (0.0%) |

#### Supplementary Table S2. Principal component analysis of dose–volume histograms

Principal component analysis (PCA) results for rectum, rectal wall, bladder and bladder wall DVHs, including eigenvalues, explained variance and dose levels associated with maximum component loadings.

| **Organ / Structure** | **Principal Component** | **Eigenvalue (SS loadings)** | **Explained variance (%)** | **Cumulative variance (%)** | **Dose at max loading (Gy)** |
| --- | --- | --- | --- | --- | --- |
| **Rectal wall** | PC1 | 23.907 | 30.65 | 30.65 | 13.0 |
|  | PC2 | 23.317 | 29.89 | 60.54 | 28.5 |
|  | PC3 | 13.026 | 16.70 | 77.24 | 3.0 |
|  | PC4 | 7.449 | 9.55 | 86.79 | 34.5 |
|  | PC5 | 4.360 | 5.59 | 92.38 | 37.5 |
| **Rectum (whole)** | PC1 | 43.665 | 44.11 | 44.11 | 42.5 |
|  | PC2 | 31.493 | 31.81 | 75.92 | 13.0 |
|  | PC3 | 10.740 | 10.85 | 86.77 | 29.0 |
|  | PC4 | 9.812 | 9.91 | 96.68 | 1.5 |
| **Bladder wall** | PC1 | 38.198 | 47.75 | 47.75 | 27.0 |
|  | PC2 | 28.450 | 35.56 | 83.31 | 3.0 |
|  | PC3 | 6.529 | 8.16 | 91.47 | 37.0 |
|  | PC4 | 3.948 | 4.94 | 96.41 | 39.0 |
| **Bladder (whole)** | PC1 | 43.95 | 50.51 | 50.51 | 6.5 |
|  | PC2 | 26.46 | 30.42 | 80.93 | 34.5 |
|  | PC3 | 13.14 | 15.10 | 96.03 | 39.5 |

#### Supplementary Table S3a. Spearman correlation between rectal DVH metrics and PTV volume

Spearman correlation coefficients (ρ) between selected rectal dose–volume histogram (DVH) metrics and planning target volume (PTV).
Only metrics identified as relevant by principal component analysis are included.

| **Rectal DVH metric** | **ρ (Spearman)** | **p-value** |
| --- | --- | --- |
| V1.5 Gy | 0.520 | < 0.001 |
| V13 Gy | 0.371 | < 0.001 |
| V29 Gy | 0.271 | < 0.001 |
| V42.5 Gy | 0.113 | 0.139 |

Correlations were assessed using Spearman’s rank correlation coefficient.

DVH metrics correspond to rectal volume receiving at least the specified dose level.

Statistical significance is reported without adjustment for multiple comparisons, as analyses are exploratory.

Selected low-to-intermediate rectal metrics (V1.5, V13 and V29 Gy) showed moderate correlations with PTV volume, whereas high-dose exposure (V42.5 Gy) demonstrated weak and non-significant correlation.

#### Supplementary Table S3b. Spearman correlation between rectal wall DVH metrics and PTV volume

Spearman correlation coefficients (ρ) between selected rectal wall dose–volume histogram (DVH) metrics and planning target volume (PTV).
Metrics were selected based on principal component analysis.

| **Rectal wall DVH metric** | **ρ (Spearman)** | **p-value** |
| --- | --- | --- |
| V3.0 Gy | 0.448 | < 0.001 |
| V13 Gy | 0.503 | < 0.001 |
| V28.5 Gy | 0.380 | < 0.001 |
| V34.5 Gy | −0.149 | 0.030 |
| V37.5 Gy | −0.054 | 0.435 |

Correlations were assessed using Spearman’s rank correlation coefficient.

DVH metrics represent rectal wall volume receiving at least the specified dose level.

Statistical significance is reported without adjustment for multiple comparisons, given the exploratory nature of the analysis.

Low-to-intermediate dose metrics showed moderate correlations with PTV volume, whereas high-dose metrics demonstrated weak or inverse correlation.

#### Supplementary Table S3c. Spearman correlation between bladder DVH metrics and PTV volume.

Spearman correlation coefficients (ρ) between selected bladder dose–volume histogram (DVH) metrics and planning target volume (PTV).
Metrics were selected based on principal component analysis.

| **Bladder DVH metric** | **ρ (Spearman)** | **p-value** |
| --- | --- | --- |
| V6.5 Gy | 0.393 | < 0.001 |
| V34.5 Gy | 0.403 | < 0.001 |
| V39.5 Gy | 0.003 | 0.974 |

Correlations were assessed using Spearman’s rank correlation coefficient.

DVH metrics represent bladder volume receiving at least the specified dose level.

Statistical significance is reported without adjustment for multiple comparisons, given the exploratory nature of the analysis.

Low-to-intermediate dose metrics (V6.5 Gy and V34.5 Gy) showed moderate correlations with PTV volume, whereas high-dose exposure (V39.5 Gy) demonstrated no correlation.

#### Supplementary Table S3d. Spearman correlation between bladder wall DVH metrics and PTV volume

Spearman correlation coefficients (ρ) between selected bladder wall dose–volume histogram (DVH) metrics and planning target volume (PTV).
Metrics were selected based on principal component analysis.

| **Bladder wall DVH metric** | **ρ (Spearman)** | **p-value** |
| --- | --- | --- |
| V3.0 Gy | 0.397 | < 0.001 |
| V27 Gy | 0.383 | < 0.001 |
| V37 Gy | 0.323 | < 0.001 |
| V39 Gy | 0.021 | 0.764 |

Correlations were assessed using Spearman’s rank correlation coefficient.

DVH metrics represent bladder wall volume receiving at least the specified dose level.

Statistical significance is reported without adjustment for multiple comparisons, given the exploratory nature of the analysis.

Low-to-intermediate dose metrics showed moderate correlations with PTV volume, whereas high-dose exposure (V39 Gy) demonstrated no meaningful correlation.

#### Supplementary Table S4a. ROC analysis of rectal DVH metrics for discrimination of late rectal toxicity ≥ grade 2

Receiver operating characteristic (ROC) analysis evaluating the discriminative performance of selected rectal dose–volume histogram (DVH) metrics for late rectal toxicity ≥ grade 2 at 12 months

| **Rectal DVH metric** | **AUC** | **95% CI (lower–upper)** | **Standard error** | **Discriminative performance** | **Clinical utility** |
| --- | --- | --- | --- | --- | --- |
| V1.5 Gy | 0.658 | 0.533–0.784 | 0.064 | Poor | Limited |
| V13 Gy | 0.757 | 0.542–0.972 | 0.110 | Fair | Moderate |
| **V18.1 Gy** | **0.868** | **0.740–0.997** | **0.066** | **Good** | **High** |
| V29 Gy | 0.827 | 0.690–0.965 | 0.070 | Good | High |
| V42.5 Gy | 0.494 | 0.482–0.506 | 0.006 | No discrimination | Minimal |

ROC analyses were performed using late rectal toxicity ≥ grade 2 as the binary endpoint.

DVH metrics represent rectal volume receiving at least the specified dose level.

Area under the curve (AUC) values were interpreted according to conventional thresholds:
poor (<0.70), fair (0.70–0.80), good (>0.80).

Confidence intervals were estimated using non-parametric bootstrapping.

Metrics were selected based on principal component analysis results.

#### Supplementary Table S4b. Optimal cut-off analysis for rectal DVH metrics associated with late rectal toxicity ≥ grade 2

| Optimal Cutoff Analysis | | | | | | |
| --- | --- | --- | --- | --- | --- | --- |
| Metric | Optimal Cutoff | Youden Index | Sensitivity | Specificity | Accuracy | Clinical Recommendation |
| Rectal V1.5 Gy | 0.9538 | 0.5802 | 1.0000 | 0.5802 | 0.5952 | Moderate discriminatory performance |
| Rectal V13 Gy | 0.5671 | 0.6543 | 1.0000 | 0.6543 | 0.6667 | Strong discriminatory performance |
| Rectal V18.1 Gy | 0.3351 | 0.7531 | 1.0000 | 0.7531 | 0.7619 | Strong discriminatory performance |
| Rectal V29 Gy | 0.0883 | 0.7160 | 1.0000 | 0.7160 | 0.7262 | Strong discriminatory performance |
| Rectal V42.5 Gy | Not determinable | 0.0000 | 1.0000 | 0.0000 | 0.0357 | Limited discriminatory performance |

Optimal cut-off values derived from ROC analysis using the Youden index.
Results are exploratory and intended to support interpretation of discriminative performance rather than to define clinical dose constraints.

Optimal cut-off values should be interpreted with caution, given the limited number of toxicity events and the single-institution nature of the cohort. These thresholds are not intended as definitive planning constraints but as hypothesis-generating indicators to guide further validation.

#### Supplementary Table S4c. ROC analysis of non-discriminative DVH metrics for late toxicity endpoints

Receiver operating characteristic (ROC) analyses for DVH metrics derived from rectal wall, bladder and bladder wall structures.
Only metrics selected by principal component analysis are reported. Results are shown to ensure analytical completeness. The 37 cc absolute-volume bladder candidate is a non-dose absolute-volume candidate retained from the source output and is reported separately from dose-based V37 Gy.

| **Structure** | **DVH metric** | **AUC** | **95% CI** | **Standard error** | **Interpretation** | **Clinical utility** |
| --- | --- | --- | --- | --- | --- | --- |
| **Rectal wall** | V3 Gy | 0.656 | 0.502–0.810 | 0.079 | Poor | Limited |
|  | V13 Gy | 0.781 | 0.514–1.000 | 0.124 | Fair | Moderate |
|  | V28.5 Gy | 0.473 | 0.173–0.774 | 0.153 | No discrimination | Minimal |
|  | V34.5 Gy | 0.441 | 0.104–0.777 | 0.172 | No discrimination | Minimal |
|  | V37.5 Gy | 0.464 | 0.000–0.952 | 0.243 | No discrimination | Minimal |
| **Bladder (whole)** | V6.5 Gy | 0.488 | 0.150–0.826 | 0.173 | No discrimination | Minimal |
|  | V18.1 Gy | 0.524 | 0.051–0.997 | 0.241 | No discrimination | Minimal |
|  | V34.5 Gy | 0.589 | 0.263–0.916 | 0.167 | No discrimination | Minimal |
|  | 37 cc bladder-volume candidate | 0.298 | 0.000–0.791 | 0.202 | No discrimination | Minimal |
|  | V37 Gy | 0.363 | 0.000–0.984 | 0.251 | No discrimination | Minimal |
| **Bladder wall** | V3 Gy | 0.299 | – | – | No discrimination | Minimal |
|  | V27 Gy | 0.227 | – | – | No discrimination | Minimal |
|  | V37 Gy | 0.474 | – | – | No discrimination | Minimal |
|  | V39 Gy | 0.454 | – | – | No discrimination | Minimal |

ROC analyses were performed using late toxicity ≥ grade 2 as the binary endpoint.

DVH metrics represent organ or organ wall volume receiving at least the specified dose level.

AUC values < 0.70 were considered to indicate limited or no discriminative performance.

Optimal cut-off analyses were not performed for these metrics due to the absence of clinically meaningful discrimination.

Missing confidence intervals for bladder wall metrics reflect model instability related to the limited number of toxicity events

Although rectal wall V13 Gy showed fair discriminative performance (AUC ≈ 0.78), its wide confidence interval and lack of robustness across complementary analyses precluded further modelling or cut-off derivation.

#### Supplementary Table S5. Integrated summary of exploratory dosimetric signals for late toxicity ≥ grade 2 after prostate SBRT (12 months)

This table provides an integrated overview of exploratory dosimetric signals across rectum, rectal wall, bladder and bladder wall.

This table provides an integrated overview of the discriminative performance and model-based associations of PCA-derived DVH metrics across rectum, rectal wall, bladder and bladder wall.

Results are exploratory and intended to support interpretation rather than to define clinical constraints.

| **Structure** | **DVH metric (PCA-derived)** | **Analysis** | **Key result** | **Interpretation** |
| --- | --- | --- | --- | --- |
| **Rectum (whole)** | **V18.1 Gy** | ROC (toxicity ≥G2) | **AUC 0.868**  **(95% CI 0.740–0.997)** | Strongest exploratory discriminative signal; interpret cautiously |
|  | V29 Gy | ROC (toxicity ≥G2) | AUC 0.827 | Complementary exploratory signal with good discrimination |
|  | V18.1 Gy | Univariable logistic regression | **Scaled OR, p = 0.047** | Statistically significant association (interpret with caution due to scaling and limited events) |
| **Rectal wall** | V13 Gy | ROC (toxicity ≥G2) | AUC 0.781 | Moderate discrimination; exploratory signal (wide CI), inferior robustness vs whole-rectum metrics |
|  | Other rectal wall metrics (e.g., V3, V28.5, V34.5, V37.5) | ROC (toxicity ≥G2) | AUC < 0.70 overall | Not supported for clinical use (limited/no discrimination) |
| **Bladder (whole)** | V34.5 Gy | ROC (toxicity ≥G2) | AUC 0.589 | No clinically meaningful discrimination |
|  | Other bladder metrics (e.g., V6.5, V18.1, 37 cc absolute-volume candidate, V37) | ROC (toxicity ≥G2) | AUC ≈ 0.30–0.59 | No discriminative ability; no predictive utility demonstrated |
| **Bladder wall** | V37 Gy | ROC (toxicity ≥G2) | AUC 0.474 | No discrimination |
|  | V39 Gy | ROC (toxicity ≥G2) | AUC 0.454 | No discrimination |
|  | V27 Gy | ROC (toxicity ≥G2) | AUC 0.227 | No discrimination |
|  | V3 Gy | ROC (toxicity ≥G2) | AUC 0.299 | No discrimination |
| **Bladder wall** | Bladder wall V37 Gy | Univariable logistic regression | p = 0.893 | No demonstrable association; model non-informative |

ROC analyses used toxicity ≥ grade 2 at 12 months as the binary endpoint.

“Scaled OR” indicates the variable was entered in a scaled form (e.g., per unit increase of a proportion/standardised unit), which may yield large OR values; emphasis should be placed on directionality and statistical significance rather than magnitude.

Confidence intervals were wide for several non-rectal models, consistent with limited event counts and lack of discriminatory signal.

This synthesis table is provided for interpretability and does not replace the full ROC tables (Supplementary Tables S4a–c)

#### Supplementary Table S6a

#### Association between non-dosimetric clinical variables and late rectal toxicity (≥ grade 2 at 12 months)

| **Variable** | **Mean (SD) – G2 Yes** | **Mean (SD) – G2 No** | **OR (univariable)** | **OR (multivariable)** |
| --- | --- | --- | --- | --- |
| Rectal volume (cc) | 107.0 (57.5) | 137.6 (35.1) | 1.02 (0.99–1.05), p=0.175 | 1.00 (0.97–1.04), p=0.912 |
| Prostate volume (cc) | 46.0 (11.5) | 41.1 (18.0) | 0.99 (0.94–1.06), p=0.637 | 0.97 (0.91–1.04), p=0.338 |
| Age at diagnosis (years) | 79.3 (4.7) | 70.5 (5.9) | 0.59 (0.33–0.85), p=0.019 | 0.57 (0.20–0.92), p=0.112 |
| Baseline PSA (ng/mL) | 14.3 (8.3) | 20.4 (81.1) | 1.00 (0.99–not estimable)†, p=0.900 | 1.04 (0.98–1.36), p=0.694 |
| ISUP grade group | 3.0 (2.0) | 1.8 (0.9) | 0.40 (0.13–1.06), p=0.065 | 0.79 (0.15–5.60), p=0.779 |

Exploratory analysis of non-dosimetric clinical variables in relation to late rectal toxicity (>= grade 2) at 12 months. Odds ratios (OR) with 95% confidence intervals are shown for univariable and sparse-event multivariable logistic regression models. Given the very small number of events (n=3), multivariable estimates are numerically fragile and should be interpreted only as transparency analyses, not as reliable independent effect estimates. G2: grade >= 2 toxicity; SD: standard deviation; PSA: prostate-specific antigen; ISUP: International Society of Urological Pathology. †Upper confidence limit not estimable because of numerical instability/quasi-complete separation in the sparse-event subgroup.

#### Supplementary Table S6b

#### Association between non-dosimetric clinical variables and late genitourinary toxicity (≥ grade 2 at 12 months)

| **Variable** | **Mean (SD) – G2 Yes** | **Mean (SD) – G2 No** | **OR (univariable)** | **OR (multivariable)** |
| --- | --- | --- | --- | --- |
| Bladder volume (cc) | 213.3 (123.5) | 239.8 (127.9) | 1.00 (0.99–1.02), p=0.770 | 1.01 (0.99–1.06), p=0.349 |
| Age at diagnosis (years) | 75.0 (11.3) | 70.6 (6.1) | 0.86 (0.60–1.11), p=0.334 | 0.56 (0.13–1.01), p=0.303 |
| Prostate volume (cc) | 39.0 (8.5) | 41.4 (17.2) | 1.01 (0.95–1.14), p=0.842 | 0.87 (0.60–1.08), p=0.316 |
| Baseline PSA (ng/mL) | 9.5 (6.3) | 21.0 (80.1) | 1.03 (1.00–1.46), p=0.746 | 9.17 (1.00–2547.97), p=0.306† |
| ISUP grade group | 3.5 (2.1) | 1.9 (1.0) | 0.31 (0.05–1.03), p=0.083 | 0.00 (0.00–0.50), p=0.239† |

Exploratory analysis of non-dosimetric clinical variables in relation to late genitourinary toxicity (>= grade 2) at 12 months. Odds ratios (OR) with 95% confidence intervals are shown for univariable and sparse-event multivariable logistic regression models. Because the endpoint included only 2 events, multivariable estimates showed complete or quasi-complete separation and are not interpretable as stable effect estimates; they are retained only for transparency. G2: grade >= 2 toxicity; SD: standard deviation; PSA: prostate-specific antigen; ISUP: International Society of Urological Pathology. †Complete or quasi-complete separation detected; these estimates are numerically unstable and not interpretable as effect sizes.

#### Supplementary Table S7

#### Key findings and clinical implications

| **Domain** | **Key finding** | **Clinical implication** |
| --- | --- | --- |
| Rectal dosimetry | Intermediate rectal dose exposure is associated with ≥ G2 toxicity at 12 months, with V18.1 Gy as the most informative exploratory metric | Consider intermediate rectal dose during plan review while preserving validated high-dose constraints |
| Structure relevance | Whole-rectum metrics outperform rectal wall parameters | Global volumetric dose burden is more relevant than focal wall irradiation |
| Bladder toxicity | No isolated dosimetric or clinical signals identified | No isolated DVH signal was demonstrated; multifactorial mechanisms remain plausible but were not proven |
| Threshold interpretation | ROC-derived cut-offs are exploratory | Should not be used as prescriptive planning constraints without external validation |
| External constraints | PACE-B constraints remain appropriate safety limits | Toxicity risk may emerge below maximum constraint thresholds |
| Modelling strategy | Continuous-variable modelling is preferred in low-event cohorts to reduce overfitting risk | Use simple, transparent modelling and avoid overinterpreting sparse-event estimates |

#### Supplementary Figure S1

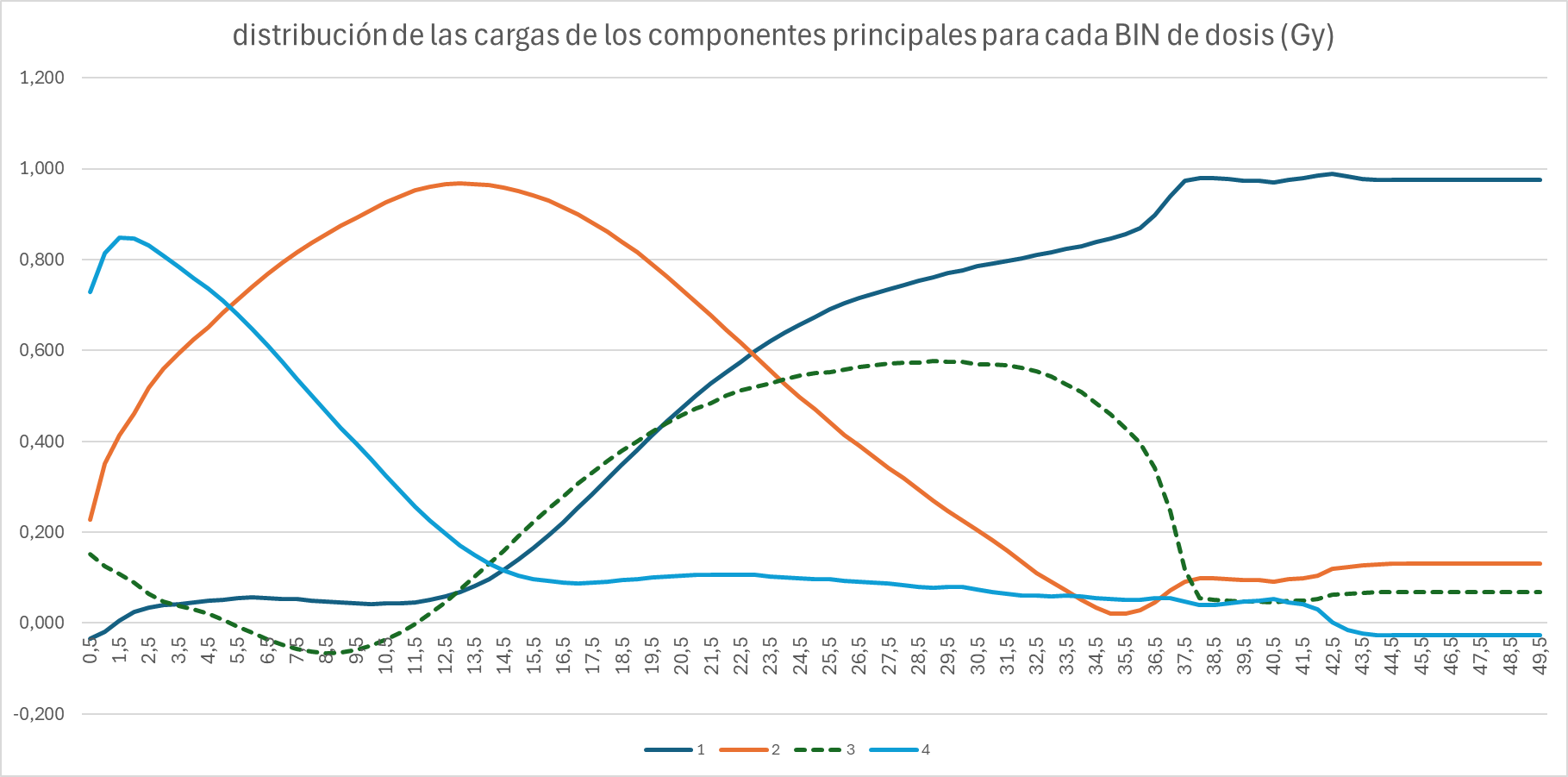

Distribution of principal component loadings across rectal dose-volume histogram dose bins.
The figure illustrates the relative contribution of each retained component to rectal DVH variability as a function of dose. For the whole rectum, the first principal component has its dominant loading in the high-dose region (42.5 Gy), whereas the second component localises to a lower-to-intermediate dose level (13.0 Gy) and the third component to 29.0 Gy. This distinction between PCA-derived variability patterns and toxicity-associated candidate metrics is discussed in the main text.

#### Supplementary Figure S2

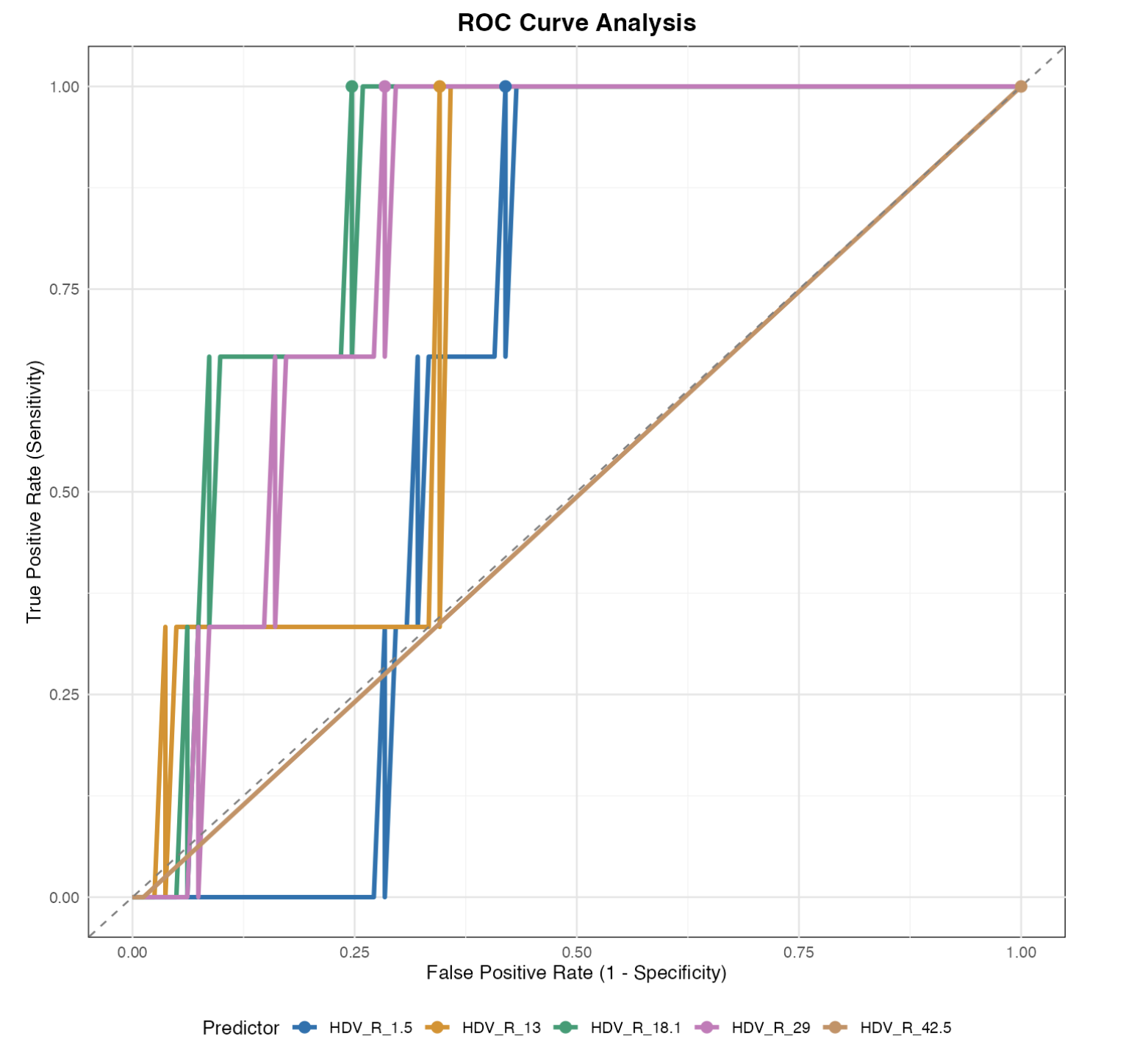

Receiver operating characteristic (ROC) curves for selected rectal dose–volume histogram metrics associated with late rectal toxicity ≥ grade 2.
Empirical ROC curves are shown for rectal V1.5 Gy, V13 Gy, V18.1 Gy, V29 Gy and V42.5 Gy. The stepwise appearance of the curves reflects the discrete nature of DVH-derived predictors and the limited number of toxicity events. Metrics corresponding to intermediate-dose exposure (V18.1 Gy and V29 Gy) demonstrate superior discriminative performance compared with low- and high-dose metrics.

#### Supplementary Figure S3

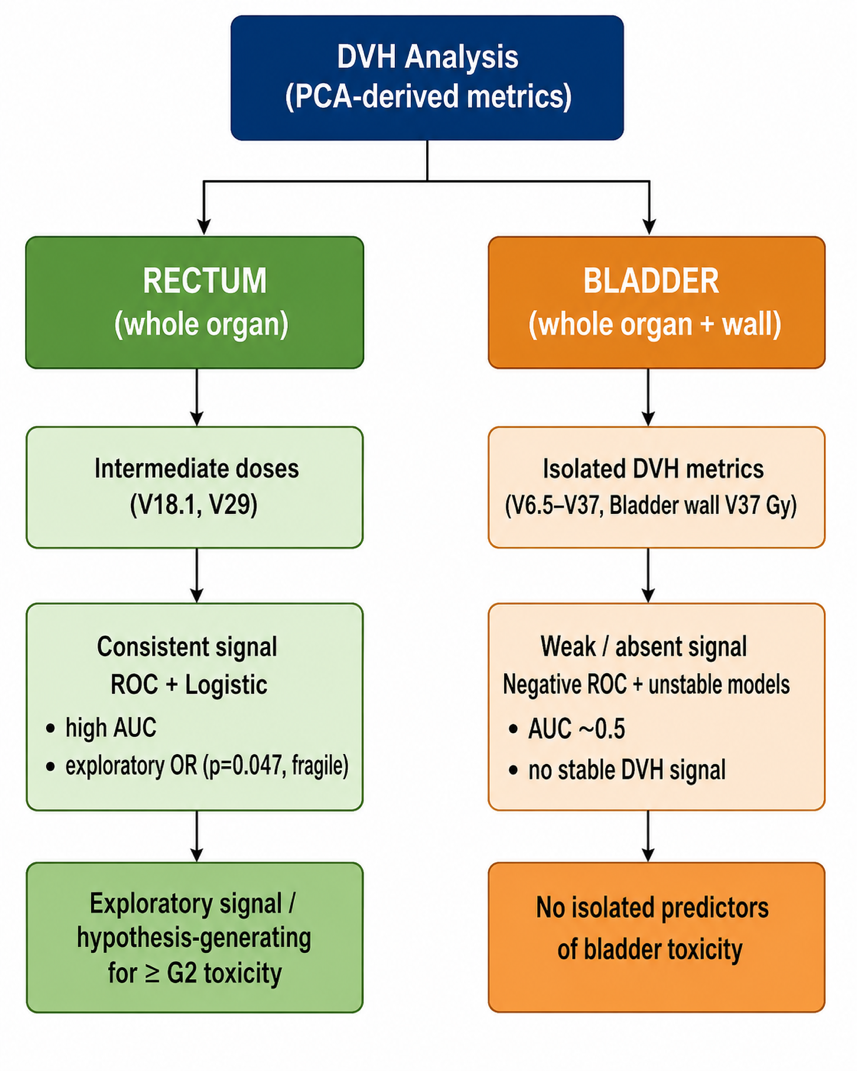

Conceptual summary of PCA-derived DVH findings for ≥ grade 2 toxicity at 12 months after prostate SBRT.

The diagram contrasts the exploratory rectal signal with the weak or absent bladder signal observed in the DVH analyses. For the whole rectum, intermediate-dose metrics (V18.1 Gy and V29 Gy) showed the most consistent hypothesis-generating association with ≥ grade 2 toxicity across ROC analysis and deliberately limited logistic regression, although the odds-ratio estimate remained fragile because of the small number of events. For bladder and bladder-wall metrics, isolated PCA-derived DVH candidates showed no stable discriminatory signal, with ROC performance close to chance and unstable regression estimates; therefore, no isolated predictor of bladder toxicity was identified.

#### Supplementary Table S8a. Patient-level phenotype of grade >=2 toxicity events in the source clinical dataset

#### Note: S8a lists patient-level grade >=2 phenotypes observed in the source clinical dataset; S8b summarises phenotype counts by follow-up time point.

Grade >=2 toxicity phenotypes were extracted from the clinical toxicity spreadsheet and cross-checked against the DVH-linked dataset. The source clinical file contains 3 GU patients with grade >=2 toxicity at 12 months, but one patient (ID-200; urinary frequency grade 2) had no valid DVH data and was therefore excluded from the DVH-linked dosimetric cohort. Accordingly, the dosimetric cohort includes 2 GU grade >=2 patients at 12 months (ID-011 and ID-231) and 3 GI grade >=2 patients (ID-179, ID-192 and ID-231).

| Patient ID | Domain | Phenotype | Grade |
| --- | --- | --- | --- |
| ID-179 | GI | constipation | 2 |
| ID-192 | GI | rectal bleeding | 2 |
| ID-231 | GI | rectal bleeding | 2 |
| ID-011 | GU | urinary urgency | 2 |
| ID-200 | GU | urinary frequency | 2 |
| ID-231 | GU | urinary tract obstruction | 2 |

Longitudinal summary of grade >=2 toxicity phenotypes. The grade 4 rectal bleeding event recorded at 18 months corresponds to ID-057 in the source clinical dataset; this patient lacked valid rectal, rectal-wall, bladder and bladder-wall DVH data in the grouped DVH file and was excluded from DVH-linked dosimetric analyses.

### Supplementary Table S8b. Time-point and phenotype-level summary of grade >=2 toxicity events in the source clinical dataset

| Month | Domain | Phenotype | Grade | n |
| --- | --- | --- | --- | --- |
| 3 | GU | urinary frequency | 2 | 2 |
| 3 | GU | urinary incontinence | 2 | 1 |
| 3 | GU | urinary tract obstruction | 2 | 1 |
| 3 | GU | urinary urgency | 2 | 1 |
| 6 | GI | rectal bleeding | 2 | 2 |
| 6 | GU | haematuria | 2 | 1 |
| 6 | GU | urinary frequency | 2 | 1 |
| 6 | GU | urinary incontinence | 2 | 1 |
| 6 | GU | urinary tract obstruction | 2 | 3 |
| 12 | GI | constipation | 2 | 1 |
| 12 | GI | rectal bleeding | 2 | 2 |
| 12 | GU | urinary frequency | 2 | 1 |
| 12 | GU | urinary tract obstruction | 2 | 1 |
| 12 | GU | urinary urgency | 2 | 1 |
| 18 | GI | rectal bleeding | 4 | 1 |
| 18 | GU | haematuria | 2 | 1 |
| 18 | GU | urinary tract obstruction | 2 | 1 |
| 18 | GU | urinary urgency | 2 | 1 |
| 24 | GU | urinary urgency | 2 | 5 |
| 30 | GU | urinary tract obstruction | 2 | 1 |
| 36 | GU | urinary tract obstruction | 2 | 1 |
| 42 | GU | haematuria | 2 | 1 |
| 42 | GU | urinary tract obstruction | 2 | 1 |

#### Supplementary Table S9. Twelve-month toxicity and DVH linkage check

This table identifies the 12-month grade >=2 toxicity events from the grouped toxicity/DVH dataset and documents whether the corresponding DVH data were valid for dosimetric analysis. The main dosimetric analysis includes two GU events with valid linked DVH data; the additional clinical-only GU event (ID-200) is shown here because it lacks valid DVH data and is therefore excluded from DVH-linked dosimetric analyses.

Abbreviations: DVH-linked, availability of valid dose-volume histogram data linkage; R, whole rectum volume; PR, rectal wall volume; V, whole bladder volume; PV, bladder wall volume; Y, valid/yes; N, not valid/no.

| Patient ID | Domain | 12-month grouped grade | DVH rectum/rectal wall valid | DVH bladder/bladder wall valid | Interpretation |
| --- | --- | --- | --- | --- | --- |
| ID-179 | GI | 2 | Y/Y | Y/Y | Included; GI event constipation in source phenotype file |
| ID-192 | GI | 2 | Y/Y | Y/Y | Included; GI event rectal bleeding |
| ID-231 | GI and GU | 2 / 2 | Y/Y | Y/Y | Included; rectal bleeding and urinary tract obstruction |
| ID-011 | GU | 2 | Y/N | Y/N | Included for grouped bladder/vesical endpoint; urinary urgency |
| ID-200 | GU | 2 | N/N | N/N | Excluded from DVH-linked dosimetric cohort; explains the extra GU event in the clinical-only dataset |

Supplementary Table S10. Institutional SBRT planning and preparation protocol

Protocol elements added for the second revision. These data describe institutional planning, preparation and delivery procedures used to contextualise the dosimetric analysis.

| Domain | Protocol element |
| --- | --- |
| Eligibility | Histologically confirmed prostate adenocarcinoma treated under an institutional protocol allowing cT1-cT3a N0 M0 disease. |
| Exclusion criteria | Previous TURP or other prostate surgery, IPSS >=19, >cT3a disease, nodal or metastatic disease, previous pelvic radiotherapy, hip prosthesis/osteosynthesis, acute haemorrhoidal syndrome or active anal fissure. |
| Prescription | 36.25 Gy in five fractions of 7.25 Gy, delivered on alternate days over a maximum of two weeks. |
| Dose escalation/boost | Although PACE-B incorporated a simultaneous integrated boost up to 40 Gy to the CTV, this cohort was restricted to a homogeneous 36.25 Gy regimen to preserve DVH comparability; sensitivity analysis including boost schedules remains pending if dose data can be harmonised. |
| Urethra-sparing objective | Urethral planning volume prescription/objective of 32.5 Gy in five fractions. |
| PTV coverage | D98% >=95% of the prescription dose; D2% <=107%, with D2% <=110% acceptable. |
| Seminal vesicles | CTV prostate only if Roach-estimated seminal-vesicle involvement risk <15%; prostate plus proximal seminal vesicles if risk >=15%. |
| PTV margin | CTV plus 5 mm isotropic margin, reduced to 3 mm posteriorly. |
| Rectal wall constraints | V100% <5%; V90% <10% optimal (<15% acceptable); V80% <20% optimal (<25% acceptable). |
| Bladder wall constraints | V100% <10-15%; V90% <20%; V50% <50%. |
| Other OAR constraints | Femoral head D2% <=50%; penile bulb mean dose <75% of the prescription dose. |
| Rectal preparation | Low-residue diet; simethicone if meteorism; rectal micro-enema on the evening before and before CT simulation or treatment. |
| Rectal immobilisation | Endorectal balloon with standard 80 cc air filling, positioned to reproduce simulation geometry. One patient in the analytical cohort was treated without a rectal balloon, explaining the lower bound of the contoured rectal-volume range. |
| Bladder preparation | Void 30-40 minutes before appointment and drink 500 mL water; CT simulation included CH10 catheterisation and 30 cc iodinated contrast; no routine catheterisation during treatment fractions. |
| Image guidance | Intraprostatic fiducial markers and cone-beam CT were used for image guidance. When fiducials were used, two VISICOIL™ fiducial markers were inserted transrectally under ultrasound guidance at least 7 days before CT simulation, whenever feasible. |
| Supportive medication | Prophylactic alpha-blocker therapy, mainly tamsulosin, before SBRT unless contraindicated or not tolerated. |
| ADT | Risk-adapted ADT: 4-6 months for unfavourable intermediate-risk disease and 18-36 months for high-risk disease, usually 24 months. Patient-level duration counts were not tabulated because start/end dates were incomplete in a subset of patients and denominators varied according to DVH availability. |
